# Reliability and Validity of the Brief Attention and Mood Scale of 7 Items (BAMS-7): A Self-Administered, Online Assessment

**DOI:** 10.1101/2024.04.05.24305401

**Authors:** Kevin P. Madore, Allen M. Osman, Kelsey R. Kerlan, Robert J. Schafer

**Affiliations:** Lumos Labs, Inc

## Abstract

Changes in technology, regulatory guidance, and COVID-19 have spurred an explosion in online studies in the social and clinical sciences. This surge has led to a need for brief and accessible instruments that are designed and validated specifically for self-administered, online use. Addressing this opportunity, the Brief Attention and Mood Scale of 7 Items (BAMS-7) was developed and validated in five cohorts across four studies to assess real-world attention and mood in one instrument. In Study 1, an exploratory factor analysis was run on responses from an initial nine-item survey in a very large, healthy, adult sample (N=75,019, ages 18-89 years). Two brief subscales comprising seven items total were defined and further characterized: one for Attention, the other for Mood. Study 2 established convergent validity with existing questionnaires in a separate sample (N=150). Study 3 demonstrated known-groups validity of each subscale using a large sample (N=58,411) of participants reporting a lifetime diagnosis of ADHD, anxiety, or depression, alongside the healthy sample of Study 1. The Attention subscale had superior discriminability for ADHD and the Mood subscale for anxiety and depression. Study 4 applied confirmatory factor analysis to data (N=3,489) from a previously published cognitive training study that used the initial nine-item survey, finding that the Attention and Mood subscales were sensitive to the intervention (compared to an active control) to different degrees. In sum, the psychometric properties and extensive normative data set (N=75,019 healthy adults) of the BAMS-7 may make it a useful instrument in assessing real-world attention and mood.

## II. Introduction

Cognition and mood are impacted by numerous medical conditions [1, 2, 3, 4], lifestyle choices [5, 6, 7], healthy development and aging [8, 9, 10, 11], and medications or other interventions [12, 13, 14, 15]. Conditions principally defined by impaired cognition – such as ADHD or mild cognitive impairment – are often associated with concomitant changes in mood status, either directly or indirectly [16, 17, 18, 19, 20, 21, 22]. Similarly, conditions principally defined by one’s mood or emotions – such as anxiety or depression – often have a corresponding impact on cognition [23, 24, 25, 26]. Given the intimate relationship between cognition and mood, the ability to measure both in one scale may be both convenient and important.

Intersecting with the need for concurrent measurement of cognition and mood is a shifting research landscape. Due to advances in technological capabilities, changes in emphases in regulatory guidance, and lasting impacts of the COVID-19 pandemic, there has been a recent surge of online studies in the social and clinical sciences [27, 28, 29, 30]. Related to this point is a growing body of contemporary research indicating that the psychometric properties of survey instruments may indeed be impacted by factors such as mode of administration and length [31, 32, 33, 34, 35, 36, 37]. As a result, instruments that are designed and characterized specifically for online research are paramount. To collect reliable responses from large numbers of participants via their own internet-connected devices, instrument qualities like brevity and accessibility of language are likewise required. For studies using relatively brief interventions, the time interval for evaluation is also important: for example, using Broadbent’s Cognitive Failures Questionnaire (CFQ) [e.g., 38, 39, 40, 41] to evaluate cognitive failures over the past six months may not be appropriate for measuring change over a shorter period of time. Furthermore, instruments that have been normed and validated based on traditional, in-person administration may have different characteristics with at-home, self-administration on one’s own computer or smart device.

To meet the needs of the present-day landscape, and to offer a very large normative data set to the research community, we present and describe a brief, seven-item scale of real-world attention and mood: the BAMS-7. The BAMS-7 complements existing instruments in the literature by emphasizing brevity, accessibility, and measures of multiple constructs (attention and mood) within one scale in a validated online format. Given that attention and mood are correlated in healthy [42, 43, 44] and clinical populations [2, 15, 17, 19], it may be advantageous to adopt one scale with separable measures of attention and mood. The scale may be useful both as an outcome measure (i.e., a dependent variable) or covariate (i.e., an independent variable) in clinical and psychological research.

### The Initial Nine-Item Survey

In 2015, Hardy et al. [45] published the results of a large, online study evaluating an at-home, computerized cognitive training program [described also in 46]. As a secondary outcome measure, the authors created a nine-item survey of “cognitive failures and successes as well as emotional status” [p. 6; 45]. This original survey is shown in **Table 1** and consisted of two parts. In a first section of four items, participants responded to questions about the frequency of real-world cognitive failures or successes within the last month. In a second section of five items, participants responded to questions about the extent of agreement with statements relating to feelings of positive or negative mood and emotions, creativity, and concentration within the last week. Responses were on a five-point Likert scale and translated to score values of 0 to 4. Hardy et al. (2015) [45] created a composite measure – the “aggregate rating” – by averaging across items.

**Table 1.**
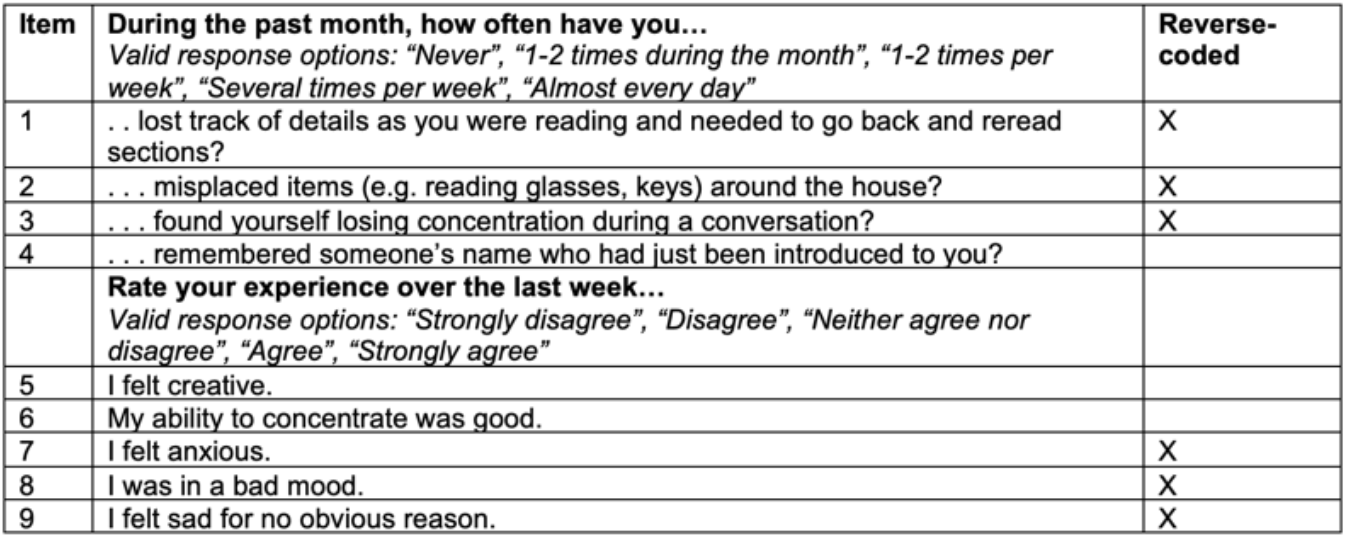
Original Hardy et al (2015) [45] Nine-Item Survey.

Although the survey was not formally characterized, the first four items were similar to ones from the CFQ, and all items had a degree of face validity. Key differences from CFQ items reflected updates for modern-day relevance (e.g. removing “newspaper” as an example item that might be misplaced around the home) and for shortening the time interval of interest to make it possible to measure changes in a shorter study. Despite the reasonable set of items, it was clear that the survey was not designed to assess a single factor or construct. Although the authors reported that the average survey rating (and several individual items) improved as a result of a cognitive training intervention, more specificity may be warranted to interpret those changes.

### The Current Research

Over the last several years, the same nine-item survey has been made available to hundreds of thousands of users of the Lumosity cognitive training program (Lumos Labs, Inc., San Francisco, CA) to inform future development of the program. Within this larger group, a subset of individuals has also provided demographic information (age, educational attainment, gender) and aspects of health history, including whether they have been diagnosed with any of a number of medical conditions. We capitalized on the availability of this massive, pre-existing data set to evaluate the original nine-item survey and to formally define and characterize a new instrument with desirable psychometric properties.

To this aim, here we present results of four studies that support sequential steps of the scale development process [see 47]. Using data from 75,019 healthy individuals in a large, online cohort in Study 1, we explore the nine-item survey used by Hardy et al. (2015) [45] and characterize a seven-item brief scale of real-world attention and mood (the BAMS-7). Through exploratory factor analysis we identify two subscales, one for Attention and one for Mood, and identify distributional and psychometric properties. In Study 2, the two subscales are shown to have convergent validity with existing questionnaires of similar constructs in a separate sample of 150 individuals from Amazon Mechanical Turk (MTurk). In Study 3, using cohorts of individuals reporting a diagnosis of ADHD (N=12,976), anxiety disorder (N=20,577), or depression (N=24,858), we show that the BAMS-7 subscales have sound known-groups validity. We demonstrate through a double dissociation that each of the two subscales (Attention and Mood) are best at discriminating between healthy controls and individuals with different clinical diagnoses; the Attention subscale is more sensitive to ADHD and the Mood subscale is more sensitive to anxiety and depression. In Study 4, we use data from the study originally reported by Hardy et al. (2015) [45] to confirm the factor structure and to evaluate sensitivity of the subscales to a cognitive training intervention.

## III. Methods

### Participants

Data from five cohorts in four studies were used in the following analyses. Survey responses from healthy participants who originally registered as members of the Lumosity cognitive training program (“Healthy cohort”) were used to develop the BAMS-7 and its subscales in Study 1. Responses from MTurk participants were used to provide convergent validity with existing questionnaires in Study 2. Responses from participants who registered through the Lumosity program and reported that they have been diagnosed with ADHD (“ADHD cohort”), anxiety disorder (“Anxiety cohort”), or depression disorder (“Depression cohort”) were used to evaluate known-groups validity of the BAMS-7 subscales in Study 3. Responses from participants in the experiment run by Hardy et al. (2015; [45]; “Hardy cohort”) were used to identify sensitivity to intervention effects in Study 4. See **Table 2** for demographic characteristics of each cohort in each study.

**Table 2.** Demographics of Each Cohort in the Current Studies.

|  | Cohort |  |  |  |  |  |
| --- | --- | --- | --- | --- | --- | --- |
|  | Healthy | MTurk | ADHD | Anxiety | Depression | Hardy |
| **N** | 75,019 | 150 | 12,976 | 20,577 | 24,858 | 3,489 |
| **Age**<br>(range, mean, SD) | 18-89yrs,<br>49.46,<br>16.32 | 20-63yrs,<br>40.16,<br>11.02 | 18-88yrs,<br>42.04,<br>15.93 | 18-89yrs,<br>47.38,<br>15.92 | 18-89yrs,<br>49.97,<br>15.87 | 18-80yrs,<br>37.73,<br>14.60 |
| **Gender**<br>(%female, %male,<br>%unknown) | 57.82%,<br>38.13%,<br>4.05% | 49.33%,<br>50.00%,<br>0.67% | 49.22%,<br>46.81%,<br>3.97% | 68.48%,<br>27.66%,<br>3.86% | 67.07%,<br>29.36%,<br>3.56% | 54.46%,<br>45.54%,<br>0% |
| **Education**<br>(%bachelor's or more,<br>%less than bachelor's,<br>%unknown) | 59.15%,<br>34.36%,<br>6.50% | 72.00%,<br>28.00%,<br>0% | 53.61%,<br>40.90%,<br>5.49% | 54.77%,<br>39.42%,<br>5.81% | 54.40%,<br>40.08%,<br>5.52% | 63.63%,<br>34.77%,<br>1.61% |
| **Study #** | 1, 3 | 2 | 3 | 3 | 3 | 4 |

Data from the Healthy, ADHD, Anxiety, and Depression cohorts were collected during normal use of a feature of the Lumosity training program. In the Lumosity Privacy Policy (www.lumosity.com/legal/privacy_policy), all participants agreed to the use and disclosure of non-personal data (e.g. de-identified or aggregate data) for any purpose. Participants were included if they were 18-89 years of age. The Healthy cohort included 75,019 participants who reported no diagnoses from a list of 34 options in an optional “information about you” survey. The ADHD cohort included 12,976 participants who reported a lifetime diagnosis of ADHD. The Anxiety cohort included 20,577 participants who reported a lifetime diagnosis of anxiety disorder. The Depression cohort included 24,858 participants who reported a lifetime diagnosis of depression disorder. Comorbid conditions were allowed in the ADHD, Anxiety, and Depression cohorts, such that a participant could be in multiple cohorts (see Supplementary Table 1 in the Supplemental Materials). Data from the additional MTurk cohort included 150 participants who were 18 or older from the general population and based in the United States without further restrictions.

The Hardy cohort included 3,489 participants who participated in the large, online, cognitive training experiment run by Hardy et al. (2015) [45] and who provided complete responses on the nine-item survey used as a secondary outcome measure. Participants ranged in age from 18-80. Individuals completed the survey prior to randomization into a cognitive training intervention group or a crossword puzzle active control group, and completed the same survey following the 10-week intervention. A complete description of the cohort and experiment can be found in Hardy et al. (2015) [45].

For the Healthy, ADHD, Anxiety, Depression, and Hardy cohorts, all available data were used, and thus the size of the population was very large. Because these were descriptive studies with pre-existing data sets, powering was not calculated; in addition, the size of each population was much larger than is commonly recommended [see 48, 49]. For the MTurk cohort, which was also a descriptive study, a power analysis indicated that 134 participants would be sufficient to detect correlations between measures with 95% power, two-tailed at *p*<.05, with expected correlation strength *r*=0.30 (G*Power 3.1) [50]. To account for potential quality issues with remote research platforms such as MTurk [28, 51, 52], our prespecified methods allowed for recruitment of up to 200 participants, with the expectation that up to 1/3 of the collected data would need to be discarded due to failed attention checks.

An institutional review board (WCG IRB; Princeton, New Jersey) provided an exempt status determination for the studies described here.

### Survey Items

Individuals in all five cohorts in the four studies took the original nine-item survey comprising items about cognitive failures, as well as mood, creativity, and concentration. As described in Hardy et al. (2015) [45] and in the Introduction, the first four survey items related to a participant’s cognitive performance over the past month, and the additional five items related to a participant’s mood and emotional status over the past week. Response options for both sets of questions were on a five-point Likert scale, but the response options differed for the two sets. Response options for the first group of questions were “Never,” “1-2 times during the month”, “1-2 times per week”, “Several times per week”, “Almost every day”. Response options for the second group of questions were “Strongly disagree”, “Disagree”, “Neither agree nor disagree”, “Agree”, “Strongly agree.”

Participants were able to skip an item by selecting “N/A” in Studies 1, 3, and 4; the MTurk sample in Study 2 did not have this option. Only participants who responded to all items were included; scores are only considered valid or complete if there are responses to all items (i.e., no N/A values).

Scoring involves numerically coding each response option on a scale from 0 to 4, where 0 represents the most negative response and 4 represents the most positive response. Items 1 (losing track of details reading), 2 (misplacing keys), 3 (losing concentration), 7 (anxious), 8 (bad mood), and 9 (sad) are reverse scored with 0 representing the most negative response and 4 representing the most positive response. Thus, with this scale, a higher item score denotes better attention or more positive mood, depending on the focus of the question.

### Concordance With Existing Questionnaires

To establish convergent validity with existing questionnaires in Study 2, data from 150 participants via MTurk were collected and analyzed for the BAMS-7. Standard attention checks were included given expected variability in the quality of MTurk participants [28, 51, 52], as detailed in the Supplemental Materials. The additional questionnaires included: the 18-item Adult ADHD Self-Report Scale with a 6-month time interval (ASRS) [53], 12-item Attention-Related Cognitive Errors Scale with an open (unspecified) time interval (ARCES) [54], 9-item Patient Health Questionnaire with a 2-week time interval (PHQ-9) [55], 20-item Positive and Negative Affect Schedule with a “few-weeks” time interval (PANAS) [56], and 7-item Generalized Anxiety Disorder questionnaire with a 2-week time interval (GAD-7) [57]. Standard scoring was adopted for each questionnaire.

#### ASRS [53]

The three outcome variables are the sum of 9 items for Part A, the sum of 9 separate items for Part B, and the sum of all 18 items for Parts A+B. Each item is rated on a five-point Likert scale (0=Never, 1=Rarely, 2=Sometimes, 3=Often, 4=Very Often). A higher sum (ranging from 0-36) for Part A denotes more inattention, a higher sum (ranging from 0-36) for Part B denotes more hyperactivity/impulsivity, and a higher sum (ranging from 0-72) for Parts A+B denotes more inattention and hyperactivity/impulsivity.

#### ARCES [54]

The outcome variable is the item mean score across the 12 items. Each item is rated on a five-point Likert scale (1=Never, 2=Rarely, 3=Sometimes, 4=Often, 5=Very Often). A higher average (ranging from 1-5) denotes more inattention.

#### PHQ-9 [55]

The outcome variable is the sum of the 9 items. Each item is rated on a four-point Likert scale (0=Not at all, 1=Several days, 2=More than half the days, 3=Nearly every day). A higher sum (ranging from 0-27) reflects greater severity of depression, where a score of 1-4 is minimal, 5-9 is mild, 10-14 is moderate, 15-19 is moderately severe, and 20-27 is severe.

#### PANAS [56]

The two outcome variables are the sum of the 10 positive items and the sum of the 10 negative items, respectively. Each item is rated on a five-point Likert scale (1=Very slightly or not at all, 2=A little, 3=Moderately, 4=Quite a bit, 5=Extremely). A higher sum (ranging from 10-50) denotes more positive affect or more negative affect, respectively.

#### GAD-7 [57]

The one outcome variable is the sum of the 7 items. Each item is rated on a four-point Likert scale (0=Not at all, 1=Several days, 2=More than half the days, 3=Nearly every day). A higher sum (ranging from 0-21) reflects greater severity of anxiety, where a score of 0-4 is minimal, 5-9 is mild, 10-14 is moderate, and 15-21 is severe.

The Supplemental Materials contain additional details regarding the MTurk design, attention checks, and questionnaires.

### Statistical Approaches

Four core approaches [58, 59, 60, 61] were implemented to establish the BAMS-7. First, exploratory factor analysis was run for the Healthy cohort in Study 1 to identify the items for the BAMS-7 and its subscales. To address collinearity and sampling distribution adequacy, correlations among the items were examined, along with the Bartlett sphericity [62] and Kaiser-Meyer-Olkin (KMO) [63] test statistics for factorability. Then, parallel analysis [64] was used to find the ideal number of latent factors to extract from the initial nine-item survey. This method is considered a superior method to Kaiser’s criterion (i.e., the Kaiser-Guttman rule) and the gold standard in factor analysis research [65, 66, 67, 68, 69]. A scree plot was also qualitatively assessed to further support the number of factors to retain. After identifying the ideal number of latent factors, the items that loaded onto each factor with a loading above 0.4 were identified following varimax rotation, maximizing item loadings on one factor and minimizing loadings on other factors. Cronbach’s alpha was utilized to determine the internal consistency of each factor.

Second, after appropriate removal of two of the nine items [70], convergent validity was established in Study 2 by examining correlations between the two subscales and existing, validated questionnaires of attention and mood in the MTurk sample. Third, known-groups validity of the BAMS-7 subscales was evaluated by conducting a receiver operating characteristic (ROC) analysis to data from ADHD, Anxiety, and Depression cohorts in Study 3. Fourth, an analysis of sensitivity to intervention effects was conducted with the Hardy cohort in Study 4, as well as a confirmatory factor analysis in this separate, independent sample. Model fit via Comparative Fit Index (CFI) and covariance between factors via Root Mean Square Error of Approximation (RMSEA) were examined for the confirmatory factor analysis [71].

### Statistical Analysis

Statistical analyses were primarily conducted in Python (version 3.9.7) using Pandas (version 1.3.5) and NumPy (version 1.20.3) and the following freely available libraries. Exploratory and confirmatory factor analysis used the factor_analyzer library (version 0.3.1). Cronbach’s alpha was computed with Pingouin (version 0.5.2), as was the ANCOVA analysis to reanalyze the original Hardy et al. (2015) [45] intervention results given the newly defined BAMS-7. Distribution skewness and kurtosis were computed with SciPy (version 1.7.3), as were correlations among questionnaires from the MTurk sample. Receiver operating characteristic (ROC) analysis used Scikit-learn (version 1.0.2). Outside of Python, JASP (version 0.17.2.1) was used to calculate CFI and RMSEA for the confirmatory factor analysis.

Unless otherwise stated, 95% confidence intervals and statistical comparisons were computed using standard bootstrap procedures [72] with 10,000 iterations.

## IV. Results

### Study 1

#### Analysis of the Original Nine-Item Survey

Survey results from the Healthy cohort (*N*=75,019) had varying degrees of inter-item (pairwise) correlation, ranging from −0.05 to 0.53, as shown in **Figure 1**. All correlations were significantly different from 0 (with bootstrapped 95% confidence intervals) at the *p*<.0001 level. Item-total correlations ranged from 0.03 (“Remembered Names”) to 0.52 (“Good Concentration”) and were all significantly different from 0 (*p*<0.0001 for all items). Cronbach’s alpha for the full survey was 0.705 (0.702-0.708). Bartlett’s test of sphericity was significant (*T*=136408.60, *p*<.0001), and the KMO statistic was acceptable at 0.77, above the commonly recommended threshold of 0.70 [67, 73]. These results indicate strong factorability and sampling adequacy in the data set.

**Figure 1.**
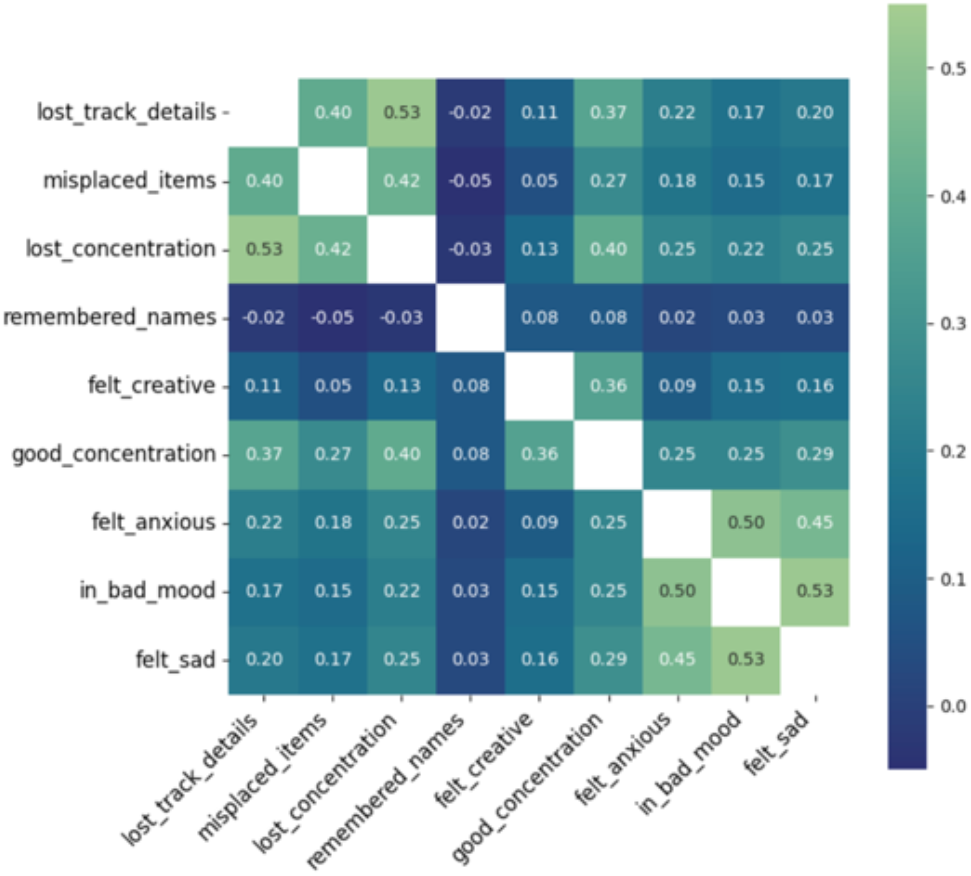
Inter-Item Correlation Coefficients of Original Nine-Item Survey in Study 1. *Note. Warmer colors show stronger correlations between respective items. All correlations are significantly different from 0 with p’s<*.*0001*.

Next, parallel analysis was used to determine the number of latent factors to retain in an exploratory factor analysis, with 41.29% of the variance explained in a resulting 3-factor solution. Additional verification by Scree plot is displayed in **Figure 2**, also indicating 3 factors clearing the eigenvalue threshold of 1.0. The initial results indicate that a 3-factor solution might be appropriate.

**Figure 2.**
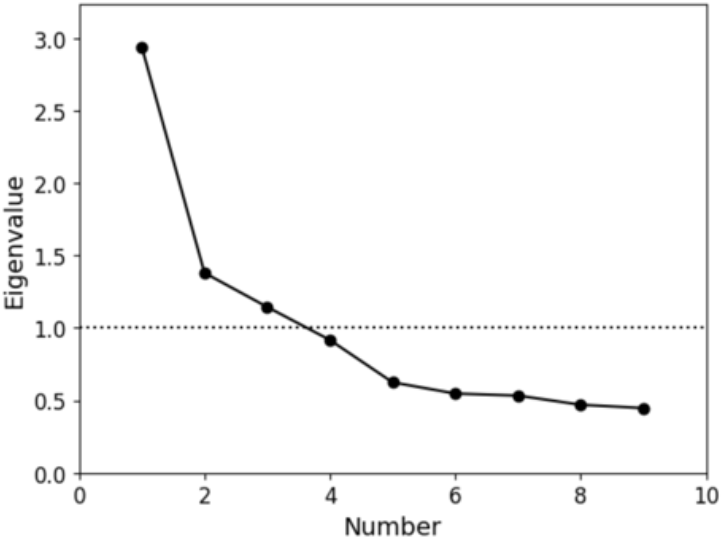
Scree Plot for Exploratory Factor Analysis of Original Nine-Item Survey in Study 1. *Note*. X-axis shows factor number and Y-axis shows eigenvalue. The dotted line 1.0 is the cutoff for inclusion in the factor solution

Then, the results of the 3-factor solution were computed using varimax rotation as shown in **Table 3**, with factor loadings of 0.4 or greater in bold type. As expected, several of the items related to cognitive successes and failures loaded together, as did several of the items related to mood. The “Remembered Names” item did not load significantly onto any of the three factors, and was dropped. The “Good Concentration” item was the only one to load strongly onto multiple factors: both the first factor, which included other items related to cognitive failures primarily associated with attention functioning, and the third factor, which included the “Felt Creative” item.

**Table 3.** Results of Exploratory Factor Analysis of Original Nine-Item Survey in Study 1.

| Item | Factor 1 | Factor 2 | Factor 3 |
| --- | --- | --- | --- |
| Lost track of details | **0.69** | 0.13 | 0.04 |
| Misplaced items | **0.56** | 0.12 | -0.04 |
| Lost concentration | **0.72** | 0.18 | 0.07 |
| Remembered names | -0.06 | 0.02 | 0.18 |
| Felt creative | 0.11 | 0.09 | **0.52** |
| Good concentration | **0.46** | 0.19 | **0.56** |
| Felt anxious | 0.19 | **0.63** | 0.05 |
| In bad mood | 0.09 | **0.77** | 0.12 |
| Felt sad | 0.16 | **0.65** | 0.16 |

*Note.* Factor loadings of 0.4 or higher are in bold type.

Cronbach’s alpha was computed to assess the internal consistency of each factor in the 3-factor solution. Factors 1 and 2 both had acceptable Cronbach’s alpha values of 0.728 and 0.745, respectively, with bootstrapped 95% confidence intervals of 0.725-0.731 and 0.742-0.748. Factor 3, however, had a lower Cronbach’s alpha value of 0.529, with a bootstrapped 95% confidence interval of 0.522-0.535. The low internal consistency and lack of an obvious description of Factor 3 led us to eliminate this factor, and subsequently to eliminate the orphaned item (“Felt Creative”) that no longer loaded onto a factor.

#### Characterization of the BAMS-7

The resulting seven-item, two-factor scale is the BAMS-7, shown in **Table 4**. On the basis of the factor analysis and the nature of the items, scores from items loading onto the first factor are averaged to compute an Attention subscale, and scores from items loading onto the second factor are averaged to compute a Mood subscale. Note that the two groups of question types in the BAMS-7 do not correspond directly to the two factors. Instead, the item on “Good Concentration,” despite falling in the second group of questions, loads onto the first factor and therefore contributes to the Attention subscale.

**Table 4.** Characterization of the BAMS-7 and its Subscales from Study 1.

| Item | During the past month, how often have you...<br>Valid response options: “Never”, “1-2 times during the month”, “1-2 times per week”, “Several times per week”, “Almost every day” | Subscale |
| --- | --- | --- |
| 1 | . . . lost track of details as you were reading and needed to go back and reread sections? | Attention |
| 2 | . . . misplaced items (e.g. reading glasses, keys) around the house? | Attention |
| 3 | . . . found yourself losing concentration during a conversation? | Attention |
|  | **Rate your experience over the last week...**<br>Valid response options: “Strongly disagree”, “Disagree”, “Neither agree nor disagree”, “Agree”, “Strongly agree” |  |
| 4 | My ability to concentrate was good. | Attention |
| 5 | I felt anxious. | Mood |
| 6 | I was in a bad mood. | Mood |
| 7 | I felt sad for no obvious reason. | Mood |

Distributional and psychometric properties of the BAMS-7 subscales are shown in **Table 5** for the Healthy cohort. Both of the subscales have modest, but statistically significant, negative skewness and kurtosis.

**Table 5.** Distributional and Psychometric Properties of the BAMS-7 in Study 1.

| Characteristics | Factor 1: Attention subscale | Factor 2: Mood subscale |
| --- | --- | --- |
| Mean | 2.3856 (2.3795, 2.3919) | 2.2359 (2.2288, 2.2428) |
| SD | 0.8598 | 0.9836 |
| Range | 0-4 | 0-4 |
| Skew | -0.4837 (-0.4955, -0.4721) | -0.1399 (-0.1508, -0.1291) |
| Kurtosis | -0.3624 (-0.3850, -0.3396) | -0.6942 (-0.7082, -0.6805) |

*Note.* Bootstrapped 95% confidence intervals are shown in parentheses as applicable. Skew and kurtosis are significantly different from 0 with $p$ 's < .0001.

Both of the subscales are related to the demographic variables of gender and age in some way. With one-way ANOVAs, the Attention subscale significantly varied with gender (*Mean*_*Male*_=2.3608, *Mean*_*Female*_=2.4011, *Mean*_*Unknown*_=2.3981; *F*=19.31, *p*<.0001) while the Mood subscale did not (*Mean*_*Male*_=0.9727, *Mean*_*Femal*e_=0.9912, *Mean*_*Unknown*_=0.9785; *F*=1.33, *p*=.27). With correlation tests, both subscales were positively associated with age within the measured range (18-89 years) (Attention: *r*=0.1994, *p*<.001; Mood: *r*=0.2285, *p*<.001). While an age-related increase on the Attention scale may be surprising given the well-established decline in cognitive performance during aging, this finding is consistent with the characteristics of the CFQ [see 41, 74; for similar results with additional questionnaires, see 75, 76]. It is also consistent with the hypothesis that self-reported cognitive failures and successes may reflect something distinct from what is measured via objective cognitive tests [77, 78].

#### Norms for the BAMS-7

A strength of the BAMS-7 is the scale of its normative data set. Normative distributions are shown across the whole population of 75,019 healthy participants for the Attention subscale (**Figure 3A**) and Mood subscale (**Figure 3D**), by gender (**Figure 3B** and **3E**), and by age in decade (**Figure 3C** and **3F**). Norm tables across the whole population, by gender, and by age in decade are also provided in look-up format for the Attention subscale (**Table 6A**) and Mood subscale (**Table 6B**).

**Table 6A.** Norm Table for the Attention Subscale across Whole Population, by Gender, and by Age in Decade.

| Population | N | 0.0 | 0.25 | 0.5 | 0.75 | 1.0 | 1.25 | 1.5 | 1.75 | 2.0 | 2.25 | 2.5 | 2.75 | 3.0 | 3.25 | 3.5 | 3.75 | 4.0 |
| --- | --- | --- | --- | --- | --- | --- | --- | --- | --- | --- | --- | --- | --- | --- | --- | --- | --- | --- |
| Healthy cohort (all genders, ages 18-89) | 75019 | 0 | 2 | 3 | 6 | 10 | 14 | 20 | 27 | 35 | 45 | 55 | 67 | 79 | 89 | 95 | 99 | 100 |
| Ages (18-29), gender f | 5059 | 1 | 3 | 6 | 11 | 16 | 22 | 29 | 38 | 47 | 58 | 68 | 79 | 88 | 94 | 98 | 100 | 100 |
| Ages (18-29), gender m | 7171 | 1 | 2 | 6 | 9 | 15 | 21 | 30 | 39 | 49 | 59 | 69 | 79 | 88 | 94 | 97 | 99 | 100 |
| Ages (30-39), gender f | 4456 | 1 | 3 | 5 | 9 | 14 | 19 | 26 | 35 | 45 | 55 | 65 | 75 | 86 | 93 | 97 | 100 | 100 |
| Ages (30-39), gender m | 4712 | 1 | 2 | 4 | 7 | 11 | 17 | 24 | 32 | 41 | 51 | 62 | 73 | 84 | 91 | 97 | 99 | 100 |
| Ages (40-49), gender f | 6650 | 1 | 2 | 4 | 8 | 12 | 17 | 23 | 30 | 39 | 49 | 59 | 70 | 82 | 91 | 97 | 99 | 100 |
| Ages (40-49), gender m | 4419 | 0 | 1 | 3 | 5 | 9 | 13 | 20 | 26 | 35 | 45 | 56 | 68 | 79 | 89 | 95 | 99 | 100 |
| Ages (50-59), gender f | 11662 | 0 | 2 | 3 | 5 | 9 | 13 | 18 | 25 | 33 | 42 | 53 | 65 | 78 | 88 | 95 | 99 | 100 |
| Ages (50-59), gender m | 5215 | 0 | 1 | 3 | 5 | 8 | 11 | 16 | 22 | 30 | 40 | 51 | 63 | 75 | 86 | 94 | 99 | 100 |
| Ages (60-69), gender f | 10931 | 0 | 1 | 2 | 4 | 6 | 9 | 14 | 19 | 26 | 35 | 46 | 59 | 72 | 84 | 94 | 98 | 100 |
| Ages (60-69), gender m | 4702 | 0 | 1 | 2 | 4 | 6 | 9 | 13 | 18 | 26 | 36 | 47 | 59 | 72 | 84 | 93 | 98 | 100 |
| Ages (70-79), gender f | 4034 | 0 | 1 | 1 | 3 | 5 | 7 | 12 | 18 | 25 | 34 | 45 | 59 | 71 | 84 | 94 | 99 | 100 |
| Ages (70-79), gender m | 2020 | 0 | 1 | 2 | 4 | 6 | 9 | 14 | 20 | 28 | 37 | 49 | 61 | 72 | 83 | 93 | 98 | 100 |
| Ages (80-89), gender f | 584 | 0 | 1 | 2 | 4 | 7 | 9 | 12 | 19 | 28 | 37 | 47 | 60 | 74 | 84 | 93 | 99 | 100 |
| Ages (80-89), gender m | 368 | 1 | 1 | 2 | 4 | 4 | 8 | 14 | 20 | 27 | 39 | 52 | 67 | 78 | 87 | 95 | 98 | 100 |

**Table 6B.** Norm Table for the Mood Subscale across Whole Population, by Gender, and by Age in Decade.

| Population | N | 0.0 | 0.33 | 0.67 | 1.0 | 1.33 | 1.67 | 2.0 | 2.33 | 2.67 | 3.0 | 3.33 | 3.67 | 4.0 |
| --- | --- | --- | --- | --- | --- | --- | --- | --- | --- | --- | --- | --- | --- | --- |
| Healthy cohort (all genders, ages 18-89) | 75019 | 2 | 4 | 8 | 16 | 24 | 35 | 47 | 58 | 69 | 82 | 89 | 95 | 100 |
| Ages (18-29), gender f | 5059 | 5 | 10 | 17 | 31 | 42 | 53 | 65 | 75 | 84 | 92 | 95 | 98 | 100 |
| Ages (18-29), gender m | 7171 | 3 | 6 | 12 | 22 | 31 | 43 | 56 | 67 | 77 | 87 | 93 | 97 | 100 |
| Ages (30-39), gender f | 4456 | 3 | 7 | 13 | 25 | 35 | 48 | 60 | 70 | 79 | 89 | 95 | 98 | 100 |
| Ages (30-39), gender m | 4712 | 2 | 5 | 10 | 19 | 29 | 41 | 54 | 65 | 75 | 87 | 93 | 97 | 100 |
| Ages (40-49), gender f | 6650 | 3 | 5 | 10 | 20 | 29 | 41 | 53 | 65 | 74 | 86 | 92 | 97 | 100 |
| Ages (40-49), gender m | 4419 | 2 | 4 | 8 | 16 | 25 | 37 | 50 | 61 | 72 | 85 | 92 | 96 | 100 |
| Ages (50-59), gender f | 11662 | 2 | 3 | 7 | 14 | 21 | 32 | 44 | 56 | 67 | 81 | 89 | 95 | 100 |
| Ages (50-59), gender m | 5215 | 1 | 3 | 6 | 13 | 22 | 32 | 44 | 56 | 67 | 80 | 89 | 95 | 100 |
| Ages (60-69), gender f | 10931 | 1 | 2 | 5 | 10 | 17 | 26 | 37 | 49 | 60 | 75 | 84 | 92 | 100 |
| Ages (60-69), gender m | 4702 | 1 | 2 | 4 | 10 | 17 | 26 | 37 | 47 | 59 | 75 | 84 | 93 | 100 |
| Ages (70-79), gender f | 4034 | 1 | 2 | 4 | 8 | 14 | 23 | 33 | 44 | 56 | 71 | 81 | 90 | 100 |
| Ages (70-79), gender m | 2020 | 1 | 2 | 4 | 8 | 14 | 21 | 32 | 42 | 55 | 69 | 80 | 91 | 100 |
| Ages (80-89), gender f | 584 | 1 | 2 | 4 | 7 | 14 | 24 | 34 | 47 | 60 | 73 | 81 | 89 | 100 |
| Ages (80-89), gender m | 368 | 0 | 1 | 2 | 7 | 14 | 23 | 35 | 46 | 57 | 71 | 82 | 92 | 100 |

*Note.* For each table, each row is a different slice of the population (or the whole population with all genders, at the top of each table). This shows the percentile (i.e., what percent of the population is less than or equal to the value shown) associated with each possible value of the respective subscale.

**Figure 3.**
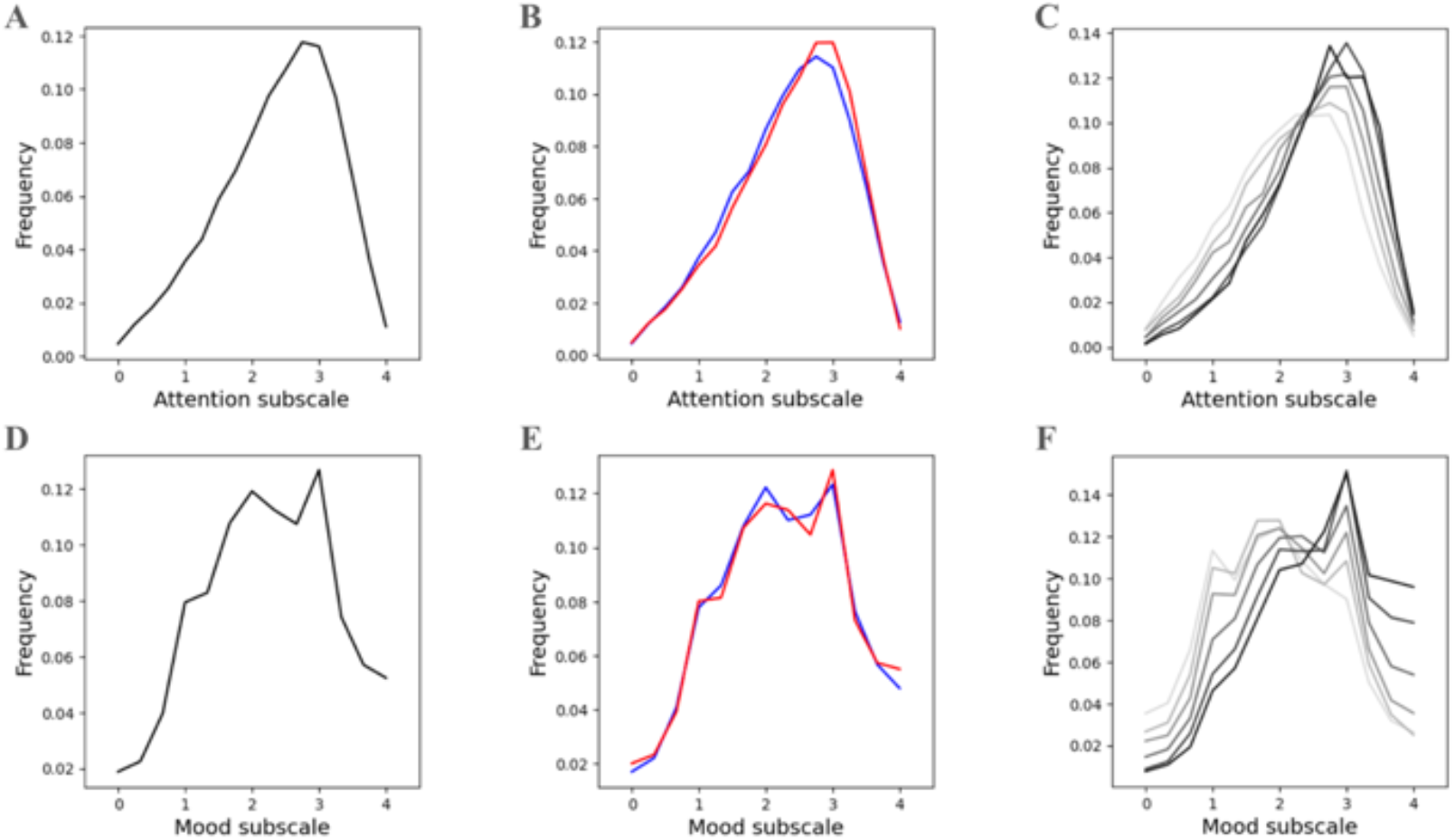
Normed Distribution of BAMS-7Attention and Mood Subscales across Whole Population, by Gender, and by Age in Decade. *Note*. Panels (A) and (D) refer to the whole normed distribution of ages and genders for Attention and Mood subscales, respectively. Panels (B) and (E) refer to distribution by gender (red=female, blue=male) across ages. Panels (C) and (F) refer to distribution by age in decade (darker color is older) across genders.

### Study 2

#### Concordance with Existing Questionnaires

To establish convergent validity, a series of correlations were computed relating the BAMS-7 Attention and Mood subscales to five known instruments of attention and mood over various timescales from the independent MTurk cohort. **Table 7A** shows *r*-values from correlations between the Attention subscale and the attention instruments, and **Table 7B** shows *r*-values for the Mood subscale with the mood instruments. All *p*-values for the correlations were < .001, meaning that the BAMS-7 Attention and Mood subscales showed significant relationships respectively with each existing questionnaire of attention and mood. The Attention subscale showed stronger relationships numerically with the attention instruments – ASRS and ARCES – while the Mood subscale showed stronger relationships with the mood instruments – GAD, PHQ, and PANAS. Note that many of the correlations are negative because a higher score on the BAMS-7 indicates better attention or mood while a higher score on each of the known instruments (excluding PANAS positive affect) indicates higher inattention or lower mood.

This pattern of results indicates that the BAMS-7 shows concordance with existing questionnaires. This can be seen in Supplementary Table 2 in the Supplemental Materials, which shows the entire set of correlations between both BAMS-7 subscales and all five existing attention and mood instruments [for similar results, see 42, 79, 80, 81]. The Supplemental Materials also contain an additional Supplementary Table 3 demonstrating strong item-level correlations between each of the BAMS-7 questions and those from the existing questionnaires with similar descriptions, demonstrating additional concordance at the item level.

**Table 7A.**
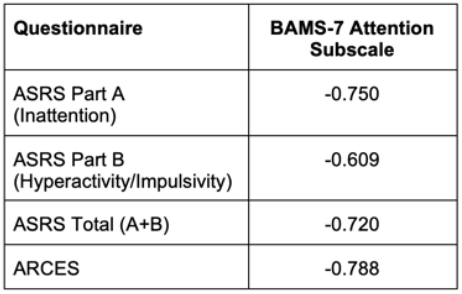
Convergent Validity of BAMS-7 Attention Subscale with Existing Attention Questionnaires in Study 2.

**Table 7B.** Convergent Validity of BAMS-7 Mood Subscale with Existing Mood Questionnaires in Study 2.

| Questionnaire | BAMS-7 Mood Subscale |
| --- | --- |
| PHQ | -0.770 |
| PANAS Pos *Aff* | 0.320 |
| PANAS Neg *Aff* | -0.780 |
| GAD | -0.765 |

Note. For each table, correlation coefficients are shown and $p's < .001$ .

### Study 3

#### Discriminatory Power of the Subscales in ADHD, Anxiety, and Depression

To evaluate the convergent and divergent validity of the BAMS-7, Attention and Mood subscale scores from the ADHD, Anxiety, and Depression cohorts were each compared to those from the Healthy cohort. A series of ROC analyses were performed to assess known-groups validity: (1) Attention subscale scores for ADHD vs Healthy, (2) Attention subscale scores for Anxiety vs Healthy, (3) Attention subscale scores for Depression vs Healthy, (4) Mood subscale scores for ADHD vs Healthy, (5) Mood subscale scores for Anxiety vs Healthy, and (6) Mood subscale scores for Depression vs Healthy. The resulting ROC curves are shown in **Figure 4A** for the Attention subscale and **4B** for the Mood subscale, and the corresponding areas under the curves (AUCs) are shown in **Table 8**.

**Table 8.** Sensitivity of BAMS-7 subscales to intervention effects using data from Hardy et al. (2015) [45].

| **AUC** | **Attention subscale** | **Mood subscale** |
| --- | --- | --- |
| ADHD vs Healthy | 0.7040 (0.6993, 0.7088) | 0.6196 (0.6144, 0.6247) |
| Anxiety vs Healthy | 0.6392 (0.6351, 0.6435) | 0.6795 (0.6755, 0.6835) |
| Depression vs Healthy | 0.6391 (0.6217, 0.6408) | 0.6587 (0.6549, 0.6626) |

*Note.* Bootstrapped 95% confidence intervals are shown in parentheses.

**Figure 4A.**
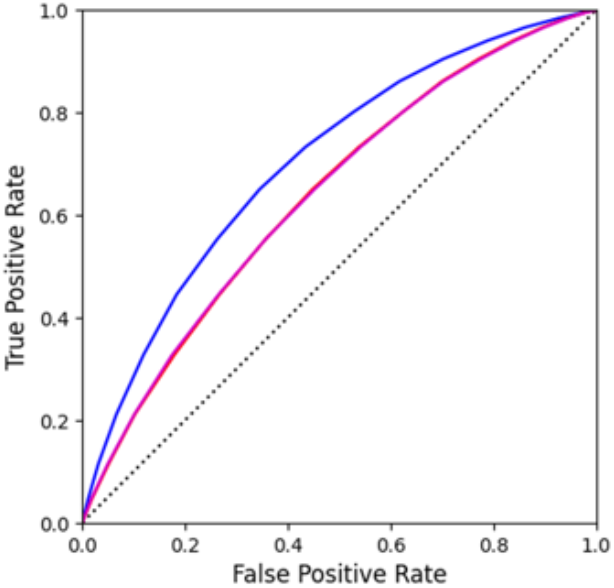
Known-groups validity evaluated with ROC Curves of BAMS-7 Attention Subscale in ADHD, Anxiety, and Depression Cohorts in Study 3.

**Figure 4B.**
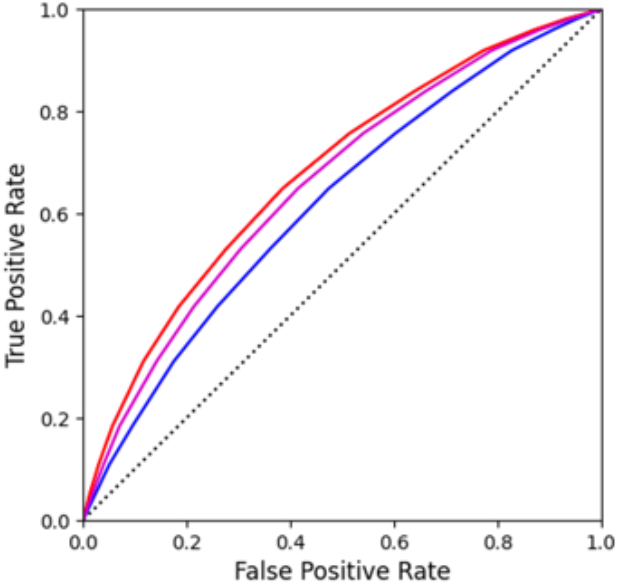
Known-groups validity evaluated with ROC Curves of BAMS-7Mood Subscale in ADHD, Anxiety, and Depression Cohorts in Study 3. *Note*. In each figure, blue=ADHD cohort, red=Anxiety cohort, and magenta=Depression cohort.

Differences within each of the three psychiatric conditions vs healthy controls were assessed by subscale to examine discriminatory ability. Within ADHD vs Healthy, the Attention subscale had a significantly higher (*p*<.0001) AUC than the Mood subscale, which provides further evidence of the factor structure of the BAMS-7, given that ADHD is primarily a disorder of attention [53]. For both the Anxiety vs Healthy and Depression vs Healthy comparisons, it was instead the Mood subscale that was significantly better (*p*<.0001) at discriminating between populations compared to the Attention subscale. This profile provides additional validation of the meaning of the subscales because mood is a hallmark of anxiety and depression [55, 57].

The ability of each of the BAMS-7 subscales to discriminate between the three psychiatric populations was assessed. Indeed, for the Attention subscale, the AUC was significantly greater (*p*<.0001) for the ADHD vs Healthy analysis relative to each of the Anxiety and Depression vs Healthy contrasts. There was no significant difference between the AUC of the Anxiety vs Healthy and Depression vs Healthy analysis for the Attention subscale. Conversely, the Mood subscale had the poorest discrimination (i.e., lowest AUC) for ADHD vs Healthy. Instead the Mood subscale had the highest (*p*<.0001) AUC for the Anxiety vs Healthy analysis, followed by Depression vs Healthy, followed by ADHD vs Healthy.

### Study 4

#### Sensitivity to a Cognitive Intervention

To confirm the factorization of the BAMS-7 and to test whether the BAMS-7 might have utility as an outcome measure in studies, we re-analyzed the data from the Hardy et al. (2015) [45] experiment using the new characterization of the BAMS-7 and its Attention and Mood subscales, excluding any participants with incomplete data. Our confirmatory factor analysis indicated adequate model fit of the BAMS-7 (CFI = 0.98 and RMSEA = 0.05), with CFI greater than 0.90 and RMSEA less than 0.08 [71]. The loadings of the seven items were also adequate and above 0.40 for the Attention subscale (item 1/rereading = 0.72, item 2/misplacing items = 0.60, item 3/losing concentration = 0.87, and item 4/good concentration = 0.48) and for the Mood subscale (item 5/anxious = 0.73, item 6/bad mood = 0.83, and item 7/sad = 0.84).

After confirming that the BAMS-7 is not sample-dependent via the confirmatory factor analysis, we followed the original analysis of Hardy et al. (2015) [45] by implementing a statistical analysis with an ANCOVA. Change in BAMS-7 subscale score (follow-up - baseline) was the dependent variable, intervention group (cognitive training or an active control) was the grouping variable, and baseline score was a covariate. Age was also included as a covariate to examine differential effects of intervention across the lifespan (for covariate results, see the Supplemental Materials).

Table 9 shows the results of the analysis of the Hardy cohort on the BAMS-7 Attention and Mood subscales. Consistent with the original analysis, there was a group (intervention) effect on the change in both the Attention and Mood subscales with the cognitive training group improving more than the active control one (Attention: *F*(1,3485)=53.73, *p*<.0001; Mood: *F*(1,3485)=17.57, *p*<.001). However, as might be expected for a cognitive intervention, the effect size (Cohen’s *d* of ANCOVA-adjusted change scores) was greater for the Attention subscale (0.247) than for the Mood subscale (0.148). These results demonstrate that the subscales of the BAMS-7 are sensitive to a cognitive intervention, and therefore may have utility as outcome measures in studies.

**Table 9.** Known-groups validity with AUC Properties of BAMS-7 Attention and Mood Subscales in ADHD, Anxiety, and Depression Cohorts in Study 3.

| **Intervention Effect** | **Active Control** | **Cognitive Training** |
| --- | --- | --- |
| Attention subscale<br>Change mean (SD) | 1.859 (2.901) | 2.409 (3.020) |
| Mood subscale<br>Change mean (SD) | 1.027 (2.811) | 1.317 (2.761) |

## V. Discussion

We describe a brief, seven-item scale of real-world attention and mood established from a very large, real-world data set: the BAMS-7. The scale is specifically designed and validated for at-home, self-administration, emphasizing brevity and accessibility—key priorities in the current research landscape—and shows potential for assessing multiple constructs effectively [31, 32, 33, 34, 35, 36, 37].

Four studies establish the validity and reliability of the BAMS-7. The scale was developed in Study 1 using data from 75,019 healthy individuals who participated in the Lumosity cognitive training program; Study 1 was also used to characterize the inter-item correlation coefficients of the original nine items of the initial survey, and to determine Cronbach’s alpha and establish distributional and psychometric properties of the BAMS-7. Concordance with existing scales for attention and mood was established in Study 2 using data from an MTurk sample. Study 3 established known-groups validity in cohorts reporting lifetime diagnoses of conditions that might be expected to have specific impairments on one or the other BAMS-7 subscale (ADHD on the Attention subscale and anxiety or depression on the Mood subscale). Study 4 re-examined data from a large-scale cognitive training study published by Hardy et al. (2015) [45] with a confirmatory factor analysis to determine whether the BAMS-7 subscales may be sensitive to cognitive interventions.

Factor analysis indicates two latent factors in the seven-item scale. Items assessing the first factor include adaptations of three items from the CFQ that focus on real-world attention function, and one item that queries the extent to which the responder agrees with the statement “I had good concentration” over the past week. The second factor includes items related to mood and anxiety. The resulting subscales – Attention and Mood, respectively – have acceptable internal consistency and descriptive statistics that may make them useful in research.

A strength of the BAMS-7 is the size and diversity of its normative data set. Age norms in the range 18-89 are provided, along with normed distribution and look-up tables across the whole population, by gender, and by age in decade for each subscale. These norms have potential to assist in comparisons from study to study and in standardized effect sizes, along with the identification of outliers [59]. Both the Attention and Mood subscales are positively correlated with age, which may appear paradoxical given the extensive literature on age-related cognitive decline. However, this relationship with age is consistent with the CFQ [e.g., 41, 74], suggesting a general divergence between objective and subjective measures of cognitive performance. It should be noted that the correlation with age is observed on a cross-sectional basis, so an alternative hypothesis is that there are generational differences in the perception of cognitive functioning. Future research should determine how subjective cognitive measures like the BAMS-7 change in longitudinal studies.

There are at least a few questions that stem from the current work on the BAMS-7. First, is it really necessary to have another psychometrically validated scale of this kind? The existing scales used in Study 2, for example, are relatively brief, validated, and commonly used in research or clinical practice. Despite the existence of other viable options, we think there remains an opportunity for instruments that are defined and validated entirely for self-administered, online use. Additionally, the scale of the available normative data set may provide advantages over existing scales for certain uses or populations. Second, is it okay that time intervals are different between attention and mood items, with the former evaluated over the past month and the latter the last week? We think that it makes sense to consider different time intervals for cognition and mood for two reasons: first, fluctuations that are significant in mood and attention may not operate on the same time scales [44, 82, 83, 84, 85]. Second, items based on the frequencies of certain behaviors – such as cognitive successes or failures – should use time intervals long enough to provide sufficient opportunities for measurement, whereas items based on a belief or emotion do not have the same requirement. The BAMS-7 includes both types of items.

A limitation of the work is that known-groups validation of the BAMS-7 may be constrained by the fact that the ADHD, Anxiety, and Depression cohorts were defined by self-reports of a lifetime clinical diagnosis. (It is worth noting, however, that population-wide studies of prevalence of clinical disorders, including ADHD, are typically based on self-report [87, 88].) Methodology for reporting diagnosis may relate to the AUC values that were obtained in Study 3: while there is no consensus threshold for adequacy of AUC values, some research has suggested that only values above 0.70 represent adequate discrimination [e.g., 86]. Most of the AUC values hovered under this cut-off. We think it is possible that the classification performance reported here is underestimated given constraints of the sample. Date of diagnosis was not reported, nor was current symptom or treatment status. If some individuals within the ADHD, Anxiety, and Depression cohorts were receiving treatment or otherwise without symptoms when taking the BAMS-7, it may be surprising that the BAMS-7 subscales could successfully discriminate at all between the three cohorts. Further use with traditionally defined cohorts, including symptom and treatment status, would be helpful.

Another limitation of the work is that the nine-item survey from which the BAMS-7 was derived was not systematically developed from an initial item pool using a deductive or inductive approach [see 47]; some of the items stem from the CFQ, but a formal item development process was not adopted for the nine-item survey itself. Instead, the development and characterization of the BAMS-7 was motivated by the pre-existence of a massive data set. Aside from the constraints of the initial pool of items, a standard and rigorous process for scale development was followed.

Overall, the pattern of results indicates that a self-administered, brief, accessible, online instrument measuring multiple constructs may be useful in the current research landscape [89, 90]. The BAMS-7 and its large normative data set show promise for improving measurement and understanding of cognition and mood in the social and clinical sciences.

## Data Availability

Aggregations of the data (i.e., norm tables) appear in the text, along with the BAMS-7 scale itself. Descriptions of the computer programming packages used to conduct the analyses are contained in the text.

## VII. Additional Notes

### Consent to participate

WCG IRB provided an exempt status for retrospective data analysis under the Code of Federal Regulations 45 CFR § 46.104(d)(4) and an exempt status for the MTurk sample under 5 CFR § 46.104(d)(2). In addition, in the Lumosity Privacy Policy (www.lumosity.com/legal/privacy_policy), all participants agreed to the use and disclosure of non-personal data (e.g. de-identified or aggregate data) for any purpose.

### Consent for publication

Not applicable.

### Availability of data and materials

Aggregations of the data (i.e., norm tables) appear in the text, along with the BAMS-7 scale itself.

### Code availability

Descriptions of the computer programming packages used to conduct the analyses are contained in the text.

### Conflicts of interest/Competing interests

This study was supported entirely by Lumos Labs, Inc. At the time of the study, KPM, AMO, KRK, and RJS were paid employees of the company, and all hold stock in the company.

Funding

No grants were received for conducting this study.

### Authors’ contributions

KPM: Conceptualization, Data Curation, Methodology, Formal Analysis, Writing – Original Draft Preparation, Writing – Review and Editing; AMO: Conceptualization, Methodology, Writing

– Review and Editing; KRK: Conceptualization, Methodology, Visualization, Writing – Review and Editing; RJS: Conceptualization, Data Curation, Methodology, Formal Analysis, Writing – Original Draft Preparation, Writing – Review and Editing.

## Acknowledgments

Lumos Labs, Inc. developed the cognitive training platform (Lumosity) and measures (BAMS-7) used in this study, as well as supported the study through the employment of KPM, AMO, KRK, and RJS. Other members of the company contributed suggestions and ideas during the design of the study and preparation of the manuscript.

## VIII. Supplemental Materials

### Study 3

**Supplementary Table 1.** Number of Participants by Cohort Condition in Study 3.

| Condition | Number of Participants |
| --- | --- |
| ADHD only | 7,057 |
| Anxiety only | 9,268 |
| Depression only | 13,325 |
| ADHD + Anxiety only | 1,769 |
| ADHD + Depression only | 1,993 |
| Anxiety + Depression only | 7,383 |
| ADHD + Anxiety + Depression | 2,157 |

### Study 2

#### MTurk Design for Concordance

We intended to recruit up to 200 individuals ages 18 and older who resided in the United States for a 7 min online Amazon MTurk HIT (human intelligence task) of “Cognition and Emotion” that paid $1.00. Various attention checks in the task reduced the sample from 200 to 150 participants for analysis (i.e., 1/4 of participants were discarded rather than our larger projection of 1/3). The attention checks involved identifying response inconsistencies, random clicking, too little or too long spent on HIT, and bots. One of the attention checks involved having participants re-rate five items from across the different questionnaires; participants were excluded if their average response mismatch was 1-point or more. After demographics, questionnaires included the ASRS, ARCES, Hardy survey (with emphasis on BAMS-7 items), PHQ-9, PANAS, and GAD-7.

#### Questionnaire Descriptions for Concordance

##### Adult ADHD Self-Report Scale (ASRS) [53]

The ASRS symptom checklist from the World Health Organization measures for probable ADHD in adults as well as ADHD symptoms. The checklist asks respondents to indicate how they have felt and conducted themselves over the past 6 months in terms of frequency of inattention and hyperactivity/impulsivity symptoms.

##### Attention-Related Cognitive Errors Scale (ARCES) [54]

The ARCES measures the frequency of cognitive errors in everyday situations that are attributed to attention lapsing. It is well-validated and reliable for remote assessment in populations across the lifespan [42, 54, 91]. Score on the ARCES is related to self-reported cognitive and clinical outcomes from independent questionnaires, including memory failures, boredom, fidgeting, mind wandering, daydreaming, media multitasking, lack of attentional control, and symptoms of depression and ADHD [79, 80, 92]. Score on the ARCES is related to task-based continuous performance test (CPT) commission errors [93], and psychometric work has shown that it is also separable from that of the CFQ [94].

##### Patient Health Questionnaire (PHQ-9) [55]

The PHQ-9 measures for probable major depressive episodes as well as depressive symptom severity. Each item represents one of the diagnostic criteria for major depressive episodes. The PHQ-9 asks participants to report the presence of each symptom within the last 2 weeks.

##### Positive and Negative Affect Schedule (PANAS) [56]

The PANAS measures positive and negative affect along various emotion and mood dimensions over various time-sensitive intervals, including over the past few weeks. The positive affect score and negative affect score are separable. It is well-validated and reliable for clinical and social sciences.

##### Generalized Anxiety Disorder (GAD-7) [57]

The GAD-7 measures for probable generalized anxiety disorder as well as anxiety symptom severity. The GAD-7 asks participants to report the presence of each symptom within the last 2 weeks.

#### MTurk Results for Concordance

**Supplementary Table 2.** Questionnaire Correlation Matrix of the BAMS-7 Attention and Mood Subscales with the Five Existing Instruments (ASRS, ARCES. PHQ-9. PANAS, and GAD-7) in Study 2.

|  | BAMS-7<br>Attention<br>subscale | BAMS-7<br>Mood<br>subscale | ASRS<br>Part A | ASRS<br>Part B | ASRS<br>Total<br>(A+B) | ARCES | PHQ-9 | PANAS<br>Pos Aff | PANAS<br>Neg Aff | GAD-7 |
| --- | --- | --- | --- | --- | --- | --- | --- | --- | --- | --- |
| BAMS-7<br>Attention<br>subscale | - |  |  |  |  |  |  |  |  |  |
| BAMS-7<br>Mood<br>subscale | 0.685*** | - |  |  |  |  |  |  |  |  |
| ASRS<br>Part A | -0.750*** | -0.659*** | - |  |  |  |  |  |  |  |
| ASRS<br>Part B | -0.609*** | -0.570*** | 0.777*** | - |  |  |  |  |  |  |
| ASRS<br>Total<br>(A+B) | -0.720*** | -0.652*** | 0.942*** | 0.943*** | - |  |  |  |  |  |
| ARCES | -0.788*** | -0.644*** | 0.834*** | 0.715*** | 0.821*** | - |  |  |  |  |
| PHQ-9 | -0.749*** | -0.770*** | 0.708*** | 0.684*** | 0.738*** | 0.718*** | - |  |  |  |
| PANAS<br>Pos Aff | 0.265** | 0.320*** | -0.118 | 0.144<sup>a</sup> | 0.014 | -0.069 | -0.178* | - |  |  |
| PANAS<br>Neg Aff | -0.685*** | -0.780*** | 0.652*** | 0.662*** | 0.697*** | 0.623*** | 0.847*** | -0.190* | - |  |
| GAD-7 | -0.755*** | -0.765*** | 0.702*** | 0.674*** | 0.730*** | 0.710*** | 0.916*** | -0.158<sup>a</sup> | 0.833*** | - |

Note. \* $p < .05$ , \*\* $p < .01$ , \*\*\* $p < .001$ , <sup>a</sup> $p < .10$ .

**Supplementary Table 3.** Item-Level Concordance Analysis with Correlation Matrix of Each BAMS-7 Item and Related Items from the Five Existing Questionnaires (ASRS. ARCES, PHQ-9, PANAS, and GAD-7) in Study 2.

| BAMS-7 | ASRS | ARCES | PANAS | PHQ-9 | GAD-7 |
| --- | --- | --- | --- | --- | --- |
| *During the last month, how often have you...* | *Over the last 6 months...* | *How frequently have these sorts of things happened to you...* | *Over the past few weeks...* | *Over the last few weeks...* | *Over the last few weeks...* |
| *Lost track of details as you were reading and needed to go back and reread sections?* | - | When reading I find that I have read several paragraphs without being able to recall what I read.<br>$r = -0.623$ | - | - | - |
| *Misplaced items (e.g., reading glasses, keys) around the house?* | How often do you misplace or have difficulty finding things at home or at work?<br>$r = -0.575$ | I have absent-mindedly misplaced frequently used objects, such as keys, pens, glasses, etc.<br>$r = -0.570$ | - | - | - |
| *Found yourself losing concentration during a conversation?* | How often do you have difficulty concentrating on what people say to you, even when they are speaking to you directly?<br>$r = -0.558$ | I have lost track of a conversation because I zoned out when someone else was talking.<br>$r = -0.676$ | - | - | - |
| *Rate your experience over the last week...* |  |  |  |  |  |
| *My ability to concentrate was good.* | - | - | Attentive<br>$r = 0.598$<br>Alert<br>$r = 0.454$ | Trouble concentrating on things, such as reading the newspaper or watching television.<br>$r = -0.403$ | - |
| *I felt anxious.* | - | - | Nervous<br>$r = 0.711$ | - | Feeling nervous, anxious, or on edge.<br>$r = -0.642$ |
| *I was in a bad mood.* | - | - | Irritable<br>$r = -0.578$ | Feeling bad about yourself - or that you are a failure or have let yourself or your family down.<br>$r = -0.470$ | - |
| *I felt sad for no obvious reason.* | - | - | Upset<br>$r = -0.627$ | Feeling down, depressed, or hopeless.<br>$r = -0.665$ | - |

Note. All $p$ 's < .0001.

### Study 4

#### ANCOVA Results with Covariates of Baseline Score and Age

In line with the original ANCOVA analysis [45], the covariate of baseline BAMS-7 on intervention effects was significant. Participants who had lower pre-intervention Attention and Mood scores exhibited greater post-intervention improvements on the Attention subscale in both Lumosity and Crosswords (*F*(1,3485)=1277.94, *p*<.001) and Mood subscale (*F*(1,3485)=1474.06, *p*<.001). There was also a significant effect of the covariate of age on the Mood subscale (*F*(1,3485)=26.57, *p*<.001) but not the Attention subscale *F*(1,3485)=0.27, *p*=.606), in Lumosity and Crosswords. Participants showed greater improvements on the Mood subscale across interventions with increasing age.

## Notes

### Author Declarations

Ethics committee/IRB of Western Institutional Review Board-Copernicus Group (WCGB IRB) gave ethical approval for this work.

### Summary of Updates

The text of the manuscript was revised to incorporate changes.

## References

1. Armstrong MJ, Okun MS. Diagnosis and treatment of Parkinson disease: a review. JAMA. 2020;323(6):548–60.

2. Bar M. A cognitive neuroscience hypothesis of mood and depression. Trends Cogn Sci. 2009;13(11):456–63.

3. Eyre H, Baune B, Lavretsky H. Clinical advances in geriatric psychiatry: a focus on prevention of mood and cognitive disorders. Psychiatr Clin. 2015;38(3):495–514.

4. Fast L, Temuulen U, Villringer K, Kufner A, Ali HF, Siebert E, et al. Machine learning-based prediction of clinical outcomes after first-ever ischemic stroke. Front Neurol. 2023;14:1114360.

5. Santos NC, Costa PS, Cunha P, Portugal-Nunes C, Amorim L, Cotter J, et al. Clinical, physical and lifestyle variables and relationship with cognition and mood in aging: a cross-sectional analysis of distinct educational groups. Front Aging Neurosci. 2014;6:21.

6. Sarris J, Thomson R, Hargraves F, Eaton M, de Manincor M, Veronese N, et al. Multiple lifestyle factors and depressed mood: a cross-sectional and longitudinal analysis of the UK Biobank (N= 84,860). BMC Med. 2020;18:1–10.

7. van Gool CH, Kempen GI, Bosma H, van Boxtel MP, Jolles J, van Eijk JT. Associations between lifestyle and depressed mood: longitudinal results from the Maastricht Aging Study. Am J Public Health. 2007;97(5):887–94.

8. Fernandes L, Wang H. Mood and cognition in old age. Front Aging Neurosci. 2018;10:286.

9. Mather M, Carstensen LL. Aging and motivated cognition: the positivity effect in attention and memory. Trends Cogn Sci. 2005;9(10):496–502.

10. Tomaszewski Farias S, De Leon FS, Gavett BE, Fletcher E, Meyer OL, Whitmer RA, et al. Associations between personality and psychological characteristics and cognitive outcomes among older adults. Psychol Aging. 2024.

11. Yurgelun-Todd D. Emotional and cognitive changes during adolescence. Curr Opin Neurobiol. 2007;17(2):251–7.

12. Keshavan MS, Vinogradov S, Rumsey J, Sherrill J, Wagner A. Cognitive training in mental disorders: update and future directions. Am J Psychiatry. 2014;171(5):510–22.

13. Koster EH, Hoorelbeke K, Onraedt T, Owens M, Derakshan N. Cognitive control interventions for depression: a systematic review of findings from training studies. Clin Psychol Rev. 2017;53:79–92.

14. Reynolds GO, Willment K, Gale SA. Mindfulness and cognitive training interventions in mild cognitive impairment: impact on cognition and mood. Am J Med. 2021;134(4):444–55.

15. Skirrow C, McLoughlin G, Kuntsi J, Asherson P. Behavioral, neurocognitive and treatment overlap between attention-deficit/hyperactivity disorder and mood instability. Expert Rev Neurother. 2009;9(4):489–503.

16. Chen C, Hu Z, Jiang Z, Zhou F. Prevalence of anxiety in patients with mild cognitive impairment: a systematic review and meta-analysis. J Affect Disord. 2018;236:211–21.

17. D’Agati E, Curatolo P, Mazzone L. Comorbidity between ADHD and anxiety disorders across the lifespan. Int J Psychiatry Clin Pract. 2019;23(4):238–44.

18. Ismail Z, Elbayoumi H, Fischer CE, Hogan DB, Millikin CP, Schweizer T, et al. Prevalence of depression in patients with mild cognitive impairment: a systematic review and meta-analysis. JAMA Psychiatry. 2017;74(1):58–67.

19. Retz W, Stieglitz RD, Corbisiero S, Retz-Junginger P, Rösler M. Emotional dysregulation in adult ADHD: what is the empirical evidence? Expert Rev Neurother. 2012;12(10):1241–51.

20. Schnyer DM, Beevers CG, Debettencourt MT, Sherman SM, Cohen JD, Norman KA, Turk-Browne NB. Neurocognitive therapeutics: from concept to application in the treatment of negative attention bias. Biology of mood & anxiety disorders. 2015;5:1–4.

21. Robison MK, Miller AL, Unsworth N. A multi-faceted approach to understanding individual differences in mind-wandering. Cognition. 2020;198:104078.

22. Yates JA, Clare L, Woods RT. Mild cognitive impairment and mood: a systematic review. Rev Clin Gerontol. 2013;23(4):317–56.

23. Gkintoni E, Ortiz PS. Neuropsychology of generalized anxiety disorder in clinical setting: a systematic evaluation. Healthcare (Basel). 2023;11(17):2446.

24. Gulpers B, Ramakers I, Hamel R, Köhler S, Voshaar RO, Verhey F. Anxiety as a predictor for cognitive decline and dementia: a systematic review and meta-analysis. Am J Geriatr Psychiatry. 2016;24(10):823–42.

25. Keller AS, Leikauf JE, Holt-Gosselin B, Staveland BR, Williams LM. Paying attention to attention in depression. Transl Psychiatry. 2019;9(1):279.

26. Williams LM. Precision psychiatry: a neural circuit taxonomy for depression and anxiety. Lancet Psychiatry. 2016;3(5):472–80.

27. Arechar AA, Rand DG. Turking in the time of COVID. Behav Res Methods. 2021;53(6):2591–5.

28. Goodman JK, Wright S. MTurk and online panel research: the impact of COVID-19, bots, TikTok, and other contemporary developments. In: Lamberton C, Rucker D, Spiller SA, editors. The cambridge handbook of consumer psychology. Cambridge: Cambridge University Press; 2022. p. 1–34.

29. Lourenco SF, Tasimi A. No participant left behind: conducting science during COVID-19. Trends Cogn Sci. 2020;24(8):583–4.

30. Saragih ID, Tonapa SI, Porta CM, Lee BO. Effects of telehealth intervention for people with dementia and their carers: a systematic review and meta-analysis of randomized controlled studies. J Nurs Scholarsh. 2022;54(6):704–19.

31. Aiyegbusi OL, Cruz Rivera S, Roydhouse J, Kamudoni P, Alder Y, Anderson N, et al. Recommendations to address respondent burden associated with patient-reported outcome assessment. Nat Med. 2024;30(3):650–9.

32. Aiyegbusi OL, Roydhouse J, Rivera SC, Kamudoni P, Schache P, Wilson R, et al. Key considerations to reduce or address respondent burden in patient-reported outcome (PRO) data collection. Nat Commun. 2022;13(1):6026.

33. Gottfried J. Practices in data-quality evaluation: a large-scale review of online survey studies published in 2022. Adv Methods Pract Psychol Sci. 2024;7(2):25152459241236414.

34. Kelfve S, Kivi M, Johansson B, Lindwall M. Going web or staying paper? the use of web-surveys among older people. BMC Med Res Methodol. 2020;20(1):252.

35. Lantos D, Moreno-Agostino D, Harris LT, Ploubidis G, Haselden L, Fitzsimons E. The performance of long vs. short questionnaire-based measures of depression, anxiety, and psychological distress among UK adults: a comparison of the patient health questionnaires, generalized anxiety disorder scales, malaise inventory, and Kessler scales. J Affect Disord. 2023;338:433–9.

36. Zager Kocjan G, Lavtar D, Sočan G. The effects of survey mode on self-reported psychological functioning: measurement invariance and latent mean comparison across face-to-face and web modes. Behav Res Methods. 2023;55(3):1226–43.

37. Wilson AB, Brooks WS, Edwards DN, Deaver J, Surd JA, Pirlo OJ, et al. Survey response rates in health sciences education research: a 10-year meta-analysis. Anat Sci Educ. 2024;17(1):11–23.

38. Bridger RS, Johnsen SÅK, Brasher K. Psychometric properties of the Cognitive Failures Questionnaire. Ergonomics. 2013;56(10):1515–24.

39. Broadbent DE, Cooper PF, FitzGerald P, Parkes KR. The Cognitive Failures Questionnaire (CFQ) and its correlates. Br J Clin Psychol. 1982;21(1):1–16.

40. Goodhew SC, Edwards M. The Cognitive Failures Questionnaire 2.0. Pers Individ Dif. 2024;218:112472.

41. Rast P, Zimprich D, Van Boxtel M, Jolles J. Factor structure and measurement invariance of the Cognitive Failures Questionnaire across the adult life span. Assessment. 2009;16(2):145–58.

42. Carriere JS, Cheyne JA, Smilek D. Everyday attention lapses and memory failures: the affective consequences of mindlessness. Conscious Cogn. 2008;17(3):835–47.

43. Hobbiss MH, Fairnie J, Jafari K, Lavie N. Attention, mindwandering, and mood. Conscious Cogn. 2019;72:1–18.

44. Irrmischer M, van der Wal CN, Mansvelder HD, Linkenkaer-Hansen K. Negative mood and mind wandering increase long-range temporal correlations in attention fluctuations. PLoS One. 2018;13(5):e0196907.

45. Hardy JL, Nelson RA, Thomason ME, Sternberg DA, Katovich K, Farzin F, et al. Enhancing cognitive abilities with comprehensive training: a large, online, randomized, active-controlled trial. PLoS One. 2015;10(9):e0134467.

46. Ng NF, Osman AM, Kerlan KR, Doraiswamy PM, Schafer RJ. Computerized cognitive training by healthy older and younger adults: age comparisons of overall efficacy and selective effects on cognition. Front Neurol. 2021;11:564317.

47. Boateng GO, Neilands TB, Frongillo EA, Melgar-Quiñonez HR, Young SL. Best practices for developing and validating scales for health, social, and behavioral research: a primer. Front Public Health. 2018;6:149.

48. Kyriazos TA. Applied psychometrics: sample size and sample power considerations in factor analysis (EFA, CFA) and SEM in general. Psychology. 2018;9(08):2207.

49. Tabachnick BG, Fidell LS. Using multivariate statistics. 6th ed. Boston: Pearson; 2013.

50. Faul F, Erdfelder E, Buchner A, Lang AG. Statistical power analyses using G* Power 3.1: tests for correlation and regression analyses. Behav Res Methods. 2009;41(4):1149–60.

51. Chmielewski M, Kucker SC. An MTurk crisis? shifts in data quality and the impact on study results. Soc Psychol Personal Sci. 2020;11(4):464–73.

52. Peer E, David R, Andrew G, Zak E, Ekaterina D. Data quality of platforms and panels for online behavioral research. Behav Res Methods. 2022;54(4):1643–62.

53. Kessler RC, Adler L, Ames M, Demler O, Faraone S, Hiripi EVA, et al. The World Health Organization Adult ADHD Self-Report Scale (ASRS): a short screening scale for use in the general population. Psychol Med. 2005;35(2):245–56.

54. Cheyne JA, Carriere JS, Smilek D. Absent-mindedness: lapses of conscious awareness and everyday cognitive failures. Conscious Cogn. 2006;15(3):578–92.

55. Kroenke K, Spitzer RL, Williams JB. The PHQ-9: Validity of a brief depression severity measure. J Gen Intern Med. 2001;16(9):606–13.

56. Watson D, Clark LA, Tellegen A. Development and validation of brief measures of positive and negative affect: the PANAS scales. J Pers Soc Psychol. 1988;54(6):1063.

57. Spitzer RL, Kroenke K, Williams JBW, Löwe B. Generalized Anxiety Disorder 7 (GAD-7). APA PsycTests. 2006.

58. Beavers AS, Lounsbury JW, Richards JK, Huck SW, Skolits GJ, Esquivel SL. Practical considerations for using exploratory factor analysis in educational research. Pract Assess Res Eval. 2013;18(1):6.

59. Brysbaert M. Designing and evaluating tasks to measure individual differences in experimental psychology: a tutorial. Cogn Res Princ Implic. 2024;9:11.

60. Costello AB, Osborne J. Best practices in exploratory factor analysis: four recommendations for getting the most from your analysis. Pract Assess Res Eval. 2005;10(1):7.

61. Knekta E, Runyon C, Eddy S. One size doesn’t fit all: using factor analysis to gather validity evidence when using surveys in your research. CBE Life Sci Educ. 2019;18(1):rm1.

62. Bartlett MS. Tests of significance in factor analysis. Br J Psychol. 1950;3:77–85.

63. Kaiser HF. An index of factorial simplicity. Psychometrika. 1974;39(1):31–6.

64. Horn JL. A rationale and test for the number of factors in factor analysis. Psychometrika. 1965;30:179–85.

65. Fabrigar LR, Wegener DT. Exploratory factor analysis. 1st ed. New York: Oxford University Press; 2011.

66. Frazier TW, Youngstrom EA. Historical increase in the number of factors measured by commercial tests of cognitive ability: are we overfactoring? Intelligence. 2007;35(2):169–82.

67. Hoelzle JB, Meyer GJ. Exploratory factor analysis: basics and beyond. In: Schinka JA, Velicer WF, Weiner IB, editors. Handbook of psychology: research methods in psychology. 2nd ed. Hoboken: John Wiley & Sons, Inc.; 2013. p. 164–88.

68. Velicer WF, Fava JL. Affects of variable and subject sampling on factor pattern recovery. Psychol Methods. 1998;3(2):231–51.

69. Watkins MW. Exploratory factor analysis: a guide to best practice. J Black Psychol. 2018;44(3):219–46.

70. Guvendir MA, Ozkan YO. Item removal strategies conducted in exploratory factor analysis: a comparative study. Int J Assess Tools Educ. 2022;9(1):165–80.

71. Browne MW, Cudeck R. Alternative ways of assessing model fit. Sociol Methods Res. 1992;21(2):230–58.

72. Wright DB, London K, Field AP. Using bootstrap estimation and the plug-in principle for clinical psychology data. J Exp Psychopathol. 2011;2(2):252–70.

73. Lloret S, Ferreres A, Hernández A, Tomás I. The exploratory factor analysis of items: guided analysis based on empirical data and software. An Psicol. 2017;33(2):417–32.

74. de Winter JC, Dodou D, Hancock PA. On the paradoxical decrease of self-reported cognitive failures with age. Ergonomics. 2015;58(9):1471–86.

75. Cyr AA, Anderson ND. Effects of question framing on self-reported memory concerns across the lifespan. Exp Aging Res. 2019;45(1):1–9.

76. Tassoni MB, Drabick DA, Giovannetti T. The frequency of self-reported memory failures is influenced by everyday context across the lifespan: implications for neuropsychology research and clinical practice. Clin Neuropsychol. 2022;37(6):1115–35.

77. Eisenberg IW, Bissett PG, Zeynep Enkavi A, Li J, MacKinnon DP, Marsch LA, et al. Uncovering the structure of self-regulation through data-driven ontology discovery. Nat Commun. 2019;10(1):1–13.

78. Yapici-Eser H, Yalcinay-Inan M, Kucuker MU, Kilciksiz CM, Yilmaz S, Dincer N, et al. Subjective cognitive assessments and n-back are not correlated, and they are differentially affected by anxiety and depression. Appl Neuropsychol Adult. 2021;1–11.

79. Franklin MS, Mrazek MD, Anderson CL, Johnston C, Smallwood J, Kingstone A, et al. Tracking distraction: the relationship between mind-wandering, meta-awareness, and ADHD symptomatology. J Atten Disord. 2017;21(6):475–86.

80. Jonkman LM, Markus CR, Franklin MS, van Dalfsen JH. Mind wandering during attention performance: effects of ADHD-inattention symptomatology, negative mood, ruminative response style and working memory capacity. PLoS One. 2017;12(7):e0181213.

81. Schubert AL, Frischkorn GT, Sadus K, Welhaf MS, Kane MJ, Rummel J. The brief mind wandering three-factor scale (BMW-3). Behavior Research Methods. 2024;56(8):8720–44.

82. Esterman M, Rothlein D. Models of sustained attention. Curr Opin Psychol. 2019;29:174–80.

83. McConville C, Cooper C. The temporal stability of mood variability. Pers Individ Dif. 1997;23(1):161–4.

84. Rosenberg MD, Scheinost D, Greene AS, Avery EW, Kwon YH, Finn ES, et al. Functional connectivity predicts changes in attention observed across minutes, days, and months. Proc Natl Acad Sci U S A. 2020;117(7):3797–807.

85. Zanesco AP, Denkova E, Jha AP. Examining long-range temporal dependence in experience sampling reports of mind wandering. Comput Brain Behav. 2022;5(2):217–33.

86. Hosmer DW, Lemeshow S. Applied logistic regression. 2nd ed. New York: Wiley; 2000.

87. Barbaresi WJ, Colligan RC, Weaver AL, Voigt RG, Killian JM, Katusic SK. Mortality, ADHD, and psychosocial adversity in adults with childhood ADHD: a prospective study. Pediatrics. 2013;131(4):637–44.

88. Faraone SV, Biederman J, Mick E. The age-dependent decline of attention deficit hyperactivity disorder: a meta-analysis of follow-up studies. Psychol Med. 2006;36(2):159–65.

89. Bowling A. Mode of questionnaire administration can have serious effects on data quality. J Public Health. 2005;27(3):281–91.

90. Rolstad S, Adler J, Rydén A. Response burden and questionnaire length: is shorter better? a review and meta-analysis. Value Health. 2011;14(8):1101–8.

## Additional References in Supplemental Materials

91. Carriere JS, Seli P, Smilek D. Wandering in both mind and body: individual differences in mind wandering and inattention predict fidgeting. Can J Exp Psychol. 2013;67(1):19–31.

92. Ralph BC, Thomson DR, Cheyne JA, Smilek D. Media multitasking and failures of attention in everyday life. Psychol Res. 2014;78(5):661–9.

93. Rosenberg M, Noonan S, DeGutis J, Esterman M. Sustaining visual attention in the face of distraction: a novel gradual-onset continuous performance task. Atten Percept Psychophys. 2013;75(3):426–39.

94. Smilek D, Carriere JS, Cheyne JA. Failures of sustained attention in life, lab, and brain: ecological validity of the SART. Neuropsychologia. 2010;48(9):2564–70.

